# The fomite contribution to the transmission of COVID-19 in the UK: an evolutionary population estimate

**DOI:** 10.1101/2021.08.11.21261903

**Authors:** A. Meiksin

## Abstract

A SEIR model with an added fomite term is used to constrain the contribution of fomites to the spread of COVID-19 under the Spring 2020 lockdown in the UK. Assuming uniform priors on the reproduction number in lockdown and the fomite transmission rate, an upper limit is found on the fomite transmission rate of less than 1 contaminated object in 7 per day per infectious person (95% CL). Basing the prior on the reproduction rate during lockdown instead on the CoMix study results for the reduction in social contacts under lockdown, and assuming the reproduction number scales with the number of social contacts, provides a much more restrictive upper limit on the transmission rate by contaminated objects of fewer than 1 in 30 per day per infectious person (95% CL). Applied to postal deliveries and groceries, the upper limit on the fomite transmission rate corresponds to a probability below 1 in 70 (95% CL) that a contaminated object transmits the infection. Fewer than about half (95% CL) of the total number of deaths during the lockdown are found to arise from fomites, and most likely fewer than a quarter. These findings apply only to fomites with a transmission rate that is unaffected by a lockdown.

## 1. Introduction

COVID-19, a respiratory infection caused by the SARS-CoV-2 virus, is believed to be transmitted primarily through viral-loaded respiratory droplets and contact with an infectious person, along with a suspected contribution from fomites (WHO, 2020). Respiratory aerosols may also be a contributing factor (Zuo et al., 2020). Establishing the role of fomites and respiratory aerosols is difficult (Lewis, 2021; Morawska and Milton, 2020). Whilst viable amounts of virus survive under laboratory conditions on contaminated surfaces (van Doremalen et al., 2020), and articles in proximity to an infectious patient can show traces of the virus RNA (Jiang et al., 2020; Ong et al., 2020), viable viruses may not survive in a natural environment in sufficient concentration to transmit the infection. On the other hand, prolonged infectivity has been measured on organic surfaces such as skin (Harbourt et al., 2020) and in a protein-rich environment (Pastorino et al., 2020), although fomites are an unlikely source of contamination in the presence of a vigorous cleaning regimen (Colaneri et al., 2020). The demonstration of indirect means of transmission in a natural environment is hampered by the expected rarity of fomites (Meiksin, 2020), so that they are difficult to detect except where an outbreak has already been identified, in which case the direction of contamination is uncertain (WHO, 2020). Indirect evidence supports the possibility of fomite transmission. Comparison between groups with and without regular hand hygiene suggests good hand hygiene reduces the transmission (Al-Ansary et al., 2020; Lio et al., 2021).

In the first half of 2020, many countries instituted national lockdowns in an effort to contain the pandemic (Flaxman et al., 2020). The decline in COVID-19 deaths following the lockdowns provides evidence that forms of non-pharmaceutical intervention that interrupt social contact successfully suppress the spread of the virus. Since a moderately high proportion of the infectious population may be pre-symptomatic or asymptomatic (WHO, 2020), even under lockdown conditions they may unwittingly contribute to the spread of COVID-19 through the accidental contamination of objects within popular circulation, such as packaged food items or mail parcels. Transmission in homes through commonly touched objects, or even through respiratory aerosols built up in poorly ventilated rooms, may also be a contributory factor.

The possibility of constraining the amount of transmission by fomites from modelling the population evolution under lockdown is explored here. It is shown that, whilst the inclusion of fomites modifies the evolution of the populations, the modification is too small to improve constraints on the rate of indirect transmission. Including the results of surveys of social contacts, however, allows more stringent constraints to be placed. On 23 March 2020, the UK Government imposed a partial lockdown, asking the public not to leave home except for essential work, outdoor exercise once per day and to buy essential items such as food and medicine. The day after the partial lockdown was imposed, the CoMix survey initiated polling of the UK adult population on their social contacts. Participants in a statistically representative sample were asked to self-report their behaviour over a 16-week period (Jarvis et al., 2020). The responses were compared to responses from a similar survey that had been conducted by the UK arm of the POLYMOD study 15 years earlier (Mossong et al., 2008). Assuming direct proportionality between the reproduction number and number of social contacts, as well as similarity in behaviour before lockdown and during the POLYMOD survey, the CoMix study inferred a reproduction number of 0.62 (95% CI: 0.37-0.89) in the UK during lockdown. Significantly, virtually all the social contacts reported were in the home, raising the question of how the epidemic, though mitigated, was able to survive well beyond the durations of the exposure and infectious periods when transmission within a household would have ceased. Evidently, transmission of the virus between households was still occurring throughout the lockdown period, possibly through non-adherence to the partial lockdown or through inter-site circulation of fomites.

In this paper, population dynamics are approximated by the SEIR model extended to include fomites to constrain the role of fomites in the epidemic in the UK. It is shown that the additional information provided by the CoMix study significantly reduces the upper limit on the fomite contribution that may be placed compared with a uniform prior on the reproduction number in lockdown, allowing for conservative modelling assumptions.

## 2. Material and methods

### 2.1. SEIR equations with a fomite term

The evolution of the pandemic is modelled using the standard set of SEIR differential equations, augmented by a fomite term (Meiksin, 2020). The equations follow the dynamics of four sub-populations: the fraction *s* of the population susceptible to infection, the fraction *e* exposed to infection, the fraction *i* of infectious individuals, and the fraction *r* of removed (recovered or perished) individuals. It is assumed here that no removed individual becomes susceptible again. Sub-populations *s* and *i* are coupled through a term *R*_*t*_*si/D*_*i*_ where *R*_*t*_, the (time-dependent) reproduction number, is the average number of people an infectious person infects. The infectious period is taken to last for a duration *D*_*i*_. The duration of an exposed individual before becoming infectious is *D*_*e*_.

A fomite term *f* represents the number of contaminated objects per capita. If *C*_*f*_ is the average number of potentially contaminatable articles a person comes into contact with per day, then *C*_*f*_ *i* is the per capita number of objects contaminated per day. (The possibility of inter-article contamination is not included.) For simplicity, an article that comes into close proximity to an infectious carrier is considered contaminated, and the average effectiveness of the contaminated article to transmit the infection is quantified through the transmittivity *T*_*f*_, representing the average number of members of the susceptible population a contaminated object infects (or, equivalently, the fraction of contaminated objects that transmit the infection to a susceptible person). The coupling term between the susceptible population and fomites is then *T*_*f*_ *sf/D*_*f*_. This represents the transmission rate per capita to an average *T*_*f*_ members of the susceptible population per capita by a number *f* of contaminated objects per capita for the duration *D*_*f*_ viruses survive on a contaminated object. The susceptible, exposed and infectious fractions depend only on the product *N*_*f*_ = *C*_*f*_ *T*_*f*_. The epidemic is initiated by the introduction of exposed and infectious carriers at the respective rates *c*_*e*_ and *c*_*i*_ per capita (of the initial population).

The model equations are

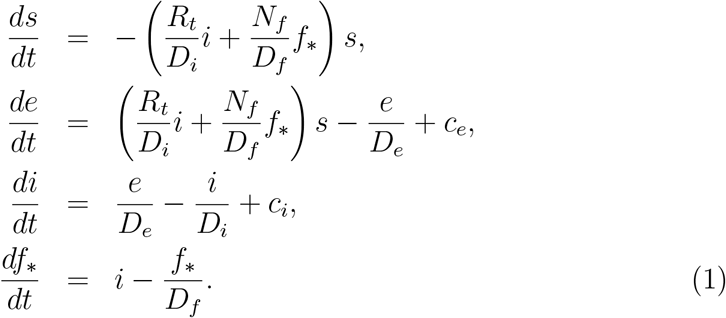

Here, the variable *f*_*_ = *f/C*_*f*_ has been introduced to explicitly show the dependence of the infectious population only on *N*_*f*_. Initially, *R*_*t*_ = *R*_0_, where *R*_0_ is the basic reproduction number when the epidemic starts. The solution to the fomite equation may be expressed in terms of the infectious population as

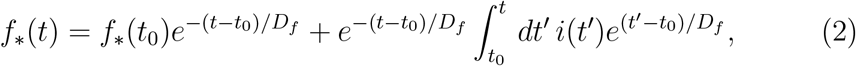

where *f*_*_(*t*_0_) is the value of *f*_*_ at an initial time *t*_0_. This form shows explicitly that the number of fomites arises from the cumulative contributions from the infectious population. In the limit *D*_*f*_≪ (*t*−*t*_0_), the fomite term is directly proportional to the instantaneous infectious population fraction when *i*(*t*) varies slowly over the time interval *D*_*f*_, *f*_*_(*t*) →*D*_*f*_ *i*(*t*). In this limit, substitution into Eqs. (1) shows that the fomites act simply to rescale the reproduction number to *R*_*t*_ + *N*_*f*_ *D*_*i*_, so that direct and indirect transmission may not be distinguished through the evolution of the populations.

The daily death rate per capita depends on the susceptible population through

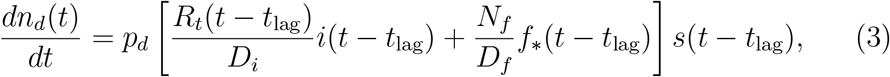

where *n*_*d*_(*t*) is the total number of deaths per capita allowing for a lag time *t*_lag_ from exposure to death and *p*_*d*_ is the fraction of infected individuals who die.

Estimates for values of the SEIR parameter are taken from (Davies et al., 2020) for COVID-19 in the UK. The initial reproduction number without intervention is estimated at *R*_0_ = 2.68±0.57. The average time from exposed to infectious state is taken to be *D*_*e*_ = 4 days, and the characteristic time during which an individual is infectious is taken to be *D*_*i*_ = 5 days. A mean infected fatality ratio 0.0050 is adopted (Meiksin, 2020; PHAS, 2020). Estimates for the mean lag time from onset of the infection to death range from 19 to 22 days (Davies et al., 2020; Flaxman et al., 2020). A time of 3 weeks is adopted here.

This paper concentrates on the lockdown period. Although *R*_*t*_ will not have changed to a new fixed value instantaneously after lockdown, for simplicity lockdown conditions are modelled by taking *R*_*t*_ = *R*_0_ before the lockdown and *R*_t,ld_ after. Estimates in the literature for both reproduction numbers generally do not distinguish direct transference of the virus from indirect transference through fomites.

The fomite contribution may in principle be identified through its effect on the evolution of the populations for finite fomite durations (*D*_*f*_ *>* 0). Laboratory estimates for the lifetime of fomites on various substances range from under an hour on copper to several hours on plastic (van Doremalen et al., 2020). When embedded in protein-rich material designed to reproduce natural concentrations, the infectivity of the virus is prolonged to as long as 4 days and possibly longer (Pastorino et al., 2020). The virus was similarly found to last at least 4 days on skin at room temperature, although its survival is severely shortened in a hot environment (Harbourt et al., 2020). Four representative duration times are considered here, 0.21 d and 0.41 d, typical of cardboard (0.14–0.30 d, 95% CI) and plastic (0.34–0.49 d, 95% CI), respectively (van Doremalen et al., 2020), 1 d and 4 d. A summary of the assumed model parameters is provided in Table 1.

**Table 1:** Model parameters. (a) Davies et al. (2020); (b) Russell et al. (2020); (c) van Doremalen et al. (2020); Pastorino et al. (2020); (d) du Plessis et al. (2020); (e) Jarvis et al. (2020)

| Parameter | Description | Value | Reference |
| --- | --- | --- | --- |
| $R_0$ | initial reproduction number | $2.1 < R_0 < 3.2$ | (a) |
| $R_{ld}$ | post-lockdown reproduction number | $0 \leq R_{t,ld} \leq 1$ | (b) |
| $T_f$ | fomite transmittivity | $0 \leq T_f \leq 0.3$ | Assumed |
| $D_e$ | duration of exposed period | 4 days | (a) |
| $D_i$ | duration of infectious period | 5 days | (a) |
| $D_f$ | duration of fomite infectious period | $0.2 \leq D_f \leq 4$ day | (c) |
| $c_0$ | peak source rate per capita | $10^{-6} \text{ day}^{-1}$ | (d) |
| $t_{c0}$ | time of source peak | day 77 | (d) |
| FWHM | source distribution full width at half maximum | 8 days | (d) |
| $\bar{f}_{ld}$ | mean reduction factor $f_{ld}$ in social contacts | 0.27 | (e) |
| $\sigma_{f_{ld}}$ | standard deviation in $f_{ld}$ | 0.086 | (e) |

An alternative means of constraining the fomite contribution is through an independent measurement of the change in the number of social contacts. Whilst a fomite contamination rate that scales in direct proportion to the number of social contacts cannot be constrained through this means, a fomite rate that is independent of, or only weakly dependent on, the mean number of social contacts may be. For example, postal deliveries and food and medical purchases continued under the lockdown, and so may have served as an indirect means of transmitting the virus between households. By contrast, indirect transmission that occurred primarily in the workplace would have been severely curtailed by the lockdown.

An estimate of the change in *R*_*t*_ before and after lockdown may be determined using the results of the CoMix survey of social contacts during lockdown (Jarvis et al., 2020). The mean number of social contacts returned from the survey was 2.9 (with an inter-quartile range of IQR = 1-4), a substantial reduction compared with the earlier POLYMOD survey result of 10.8 (IQR = 6-14). This reduction factor is adopted to provide a normal-distributed prior on *R*_*t*_ during the lockdown, taking into account a possible contribution of fomites. Here, *N*_*f*_ is assumed to apply to a fomite component that was unchanged by the lockdown, and so is held constant before and after the lockdown. If additional fomites were present, then they would have contributed even more to the overall death rate, including before the lockdown.

### 2.2. Power-series approximate solutions

Insight into the dynamical role played by fomites is provided by an approximate power-series solution to Eqs. (1). The first two equations show that the role of the fomites may be absorbed into an effective reproduction number

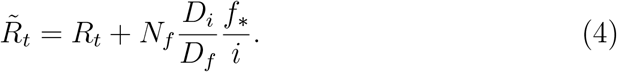

This is a formal expression, in that the evolution of both *f*_*_ and *i* depend on *R*_*t*_. Evaluating 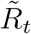 at its value near lockdown, however, provides an accurate description of the evolution of the populations afterwards. This may be demonstrated through power-series solutions to Eqs. (1). When the removed fraction *r* is small, it is convenient to use *r* as the independent variable rather than *t*. Power-series solutions around *r* = *r*_0_ = *r*(*t*_0_) after a time *t*_0_ are sought in the form:

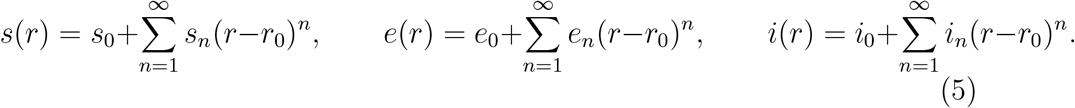

The solution to second order is described in the Appendix. The first order coefficients are

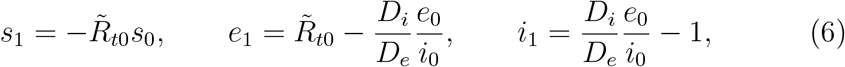

where the effective reproduction number 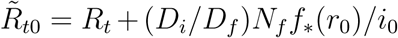 has been defined. The role of the fomites, other than in rescaling *R*_*t*_, arises only at second order (and not until even higher order for *i*), so that the presence of fomites may not be inferred from the evolution of the populations unless the fomite contribution is large and with a long duration.

The time development of the death rate, Eq. (3), is affected by fomites only at the second and higher order contributions from fomites. The role of fomites may then in principle be detected either through their second order contributions, or from independent knowledge of the reproduction number *R*_*t*_. Each of these possibilities is considered in turn.

### 2.3. Statistical quantification

Eq. (3) is used to model the mortality rates from COVID-19, seeking maximum likelihood estimates for *R*_*t*_ and *N*_*f*_. Following Flaxman et al. (2020), the number of deaths is drawn from a negative binomial distribution with mean *N*_*d*_ and variance 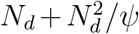, where *N*_*d*_ is the model prediction for the mean number of weekly deaths and *ψ* is a free parameter. In the limit *ψ* → ∞, the distribution becomes Poisson. Following Flaxman et al. (2020), *ψ* is drawn from a half normal distribution with mode 0 and variance 5.

The model parameters are fit using the post-lockdown data and before lockdown is eased. The general decline in mortality rate is well-modelled using a constant reproduction rate (Flaxman et al., 2020). To seek a fomite contribution, a two-step procedure is followed. First maximum likelihood values for *R*_0_ and *R*_t,ld_ are found assuming no fomites contribute (*N*_*f*_ = 0); then, for the value of *R*_0_ obtained, the joint likelihood for *R*_t,ld_ and *N*_*f*_ (≥0) is computed to determine whether adding a fomite term increases the likelihood. This procedure is followed because *R*_0_ is made uncertain by limitations in assessing the death rate during the rapid rise of the infections. This uncertainty could produce possibly spurious higher likelihoods into lockdown when a fomite term is added if pre-lockdown deaths are included in the likelihood. The limits obtained are conservative in that the procedure minimizes the fomite signal arising from terms that may not be absorbed into a re-scaling of *R*_t,ld_. Since any signal found may in principle be attributable to evolution in *R*_t,ld_ during lockdown, the signal is regarded as an upper limit to the fomite contribution.

### 2.4. Reported mortality rates

The COVID-19 mortality rates compiled by the European Centre for Disease Control ^1^ (ECDC) are used for fitting the model parameters. The ECDC compiles data from up to 500 sources in each country each week from national and regional competent authorities.

## 3. Results

### 3.1. Uniform priors

A maximum likelihood fit to the death rates from the start of lockdown on 23 March 2020 until its easing in July 2020 gives a basic reproduction number *R*_0_≃3.107 and *R*_t,ld_ = 0.760 during the lockdown period. (Although not used in this analysis, after the lockdown is eased the maximum-likelihood model for the death rates until the end of August corresponds to *R*_t,lde_ = 0.867.) The solutions with fomites show only small departures in the death rates from a model with constant reproduction rate after lockdown, in accordance with the power-series approximation (see Appendix). The magnitude of the departure depends on the duration *D*_*f*_ of the fomites. The average numbers of weekly deaths predicted are shown in the upper panel of Fig. 1, along with the 68% confidence range in uncertainty during lockdown.

**Figure 1:**
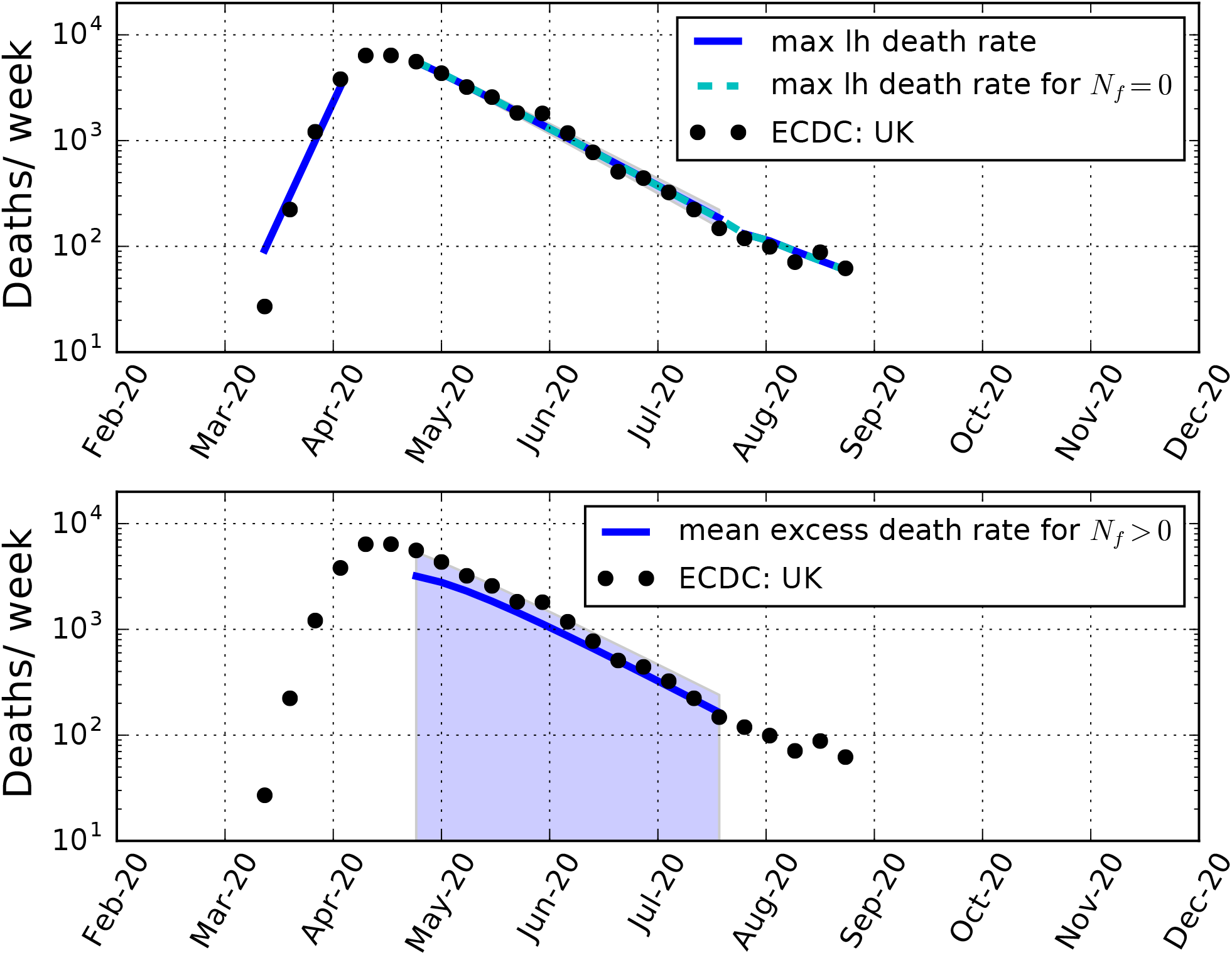
*Upper panel:* Weekly death rates in the UK. Black data points: the rates reported by the ECDC. Solid blue lines: Mean weekly deaths predicted for *D*_*f*_ = 0.41 d and assuming uniform priors for *N*_*f*_ and *R*_t,ld_ after lockdown. Also shown is the 68% confidence range of uncertainty in the mean predicted number of deaths during lockdown (shaded region). Broken cyan line: Maximum likelihood fit without a fomite contribution. *Bottom panel:* Solid blue line: Mean predicted excess deaths per week arising from fomites along with the 95% confidence upper limit (shaded region), assuming uniform priors for *N*_*f*_ and *R*_t,ld_.

The joint likelihood function for *R*_t,ld_ and *N*_*f*_ during lockdown is very flat, with the highest values corresponding to a ridge at *R*_t,ld_ + *D*_*i*_*N*_*f*_ ≈ 0.76 for 0.21 ≤ *D*_*f*_, ≤ 4 d. The probability distribution for *R*_t,ld_, marginalised over *N*_*f*_, is shown in the upper panel of Fig. 2 for *D*_*f*_ = 0.41. The results are nearly independent of *D*_*f*_ for 0.21 ≤ *D*_*f*_ ≤ 4 d. The marginalised 95% confidence upper limits on *N*_*f*_ are provided in Table 2. The maximum likelihood values for *R*_t,ld_ corresponding to the 95% confidence upper limit values for *N*_*f*_ are in the next column. Also shown are the marginalised 95% confidence upper limits on *R*_t,ld_ and the corresponding maximum likelihood values for *N*_*f*_. The upper limits on *N*_*f*_ and *R*_t,ld_ are not very sensitive to the fomite duration *D*_*f*_, and correspond to the pandemic’s being spread almost entirely by either fomites or direct transmission, respectively. The overall maximum likelihood solution corresponds to *N*_*f*_ = 0 and *R*_t,ld_ = 0.76 for all values of 0.21≤*D*_*f*_≤4 d.

**Table 2:** Model results for uniform priors in *N*_*f*_ and *R*_t,ld_. The columns are: (1) fomite duration (days); (2) 95% confidence upper limit on *N*_*f*_ (per infectious person per day) and (3) the corresponding maximum likelihood value for *R*_t,ld_; (4) 95% confidence upper limit on *R*_t,ld_ and (5) the corresponding maximum likelihood value for *N*_*f*_ (per infectious person per day).

| $D_f$ | $N_f$ (95% CL) | $R_{t,ld}$ (M-L) | $R_{t,ld}$ (95% UL) | $N_f$ (M-L) |
| --- | --- | --- | --- | --- |
| d | d<sup>-1</sup> |  |  | d<sup>-1</sup> |
| 0.21 | 0.14 | 0.04 | 0.72 | 0.013 |
| 0.41 | 0.14 | 0.04 | 0.72 | 0.013 |
| 1 | 0.14 | 0.04 | 0.72 | 0.013 |
| 4 | 0.13 | 0.05 | 0.73 | 0.009 |

**Figure 2:**
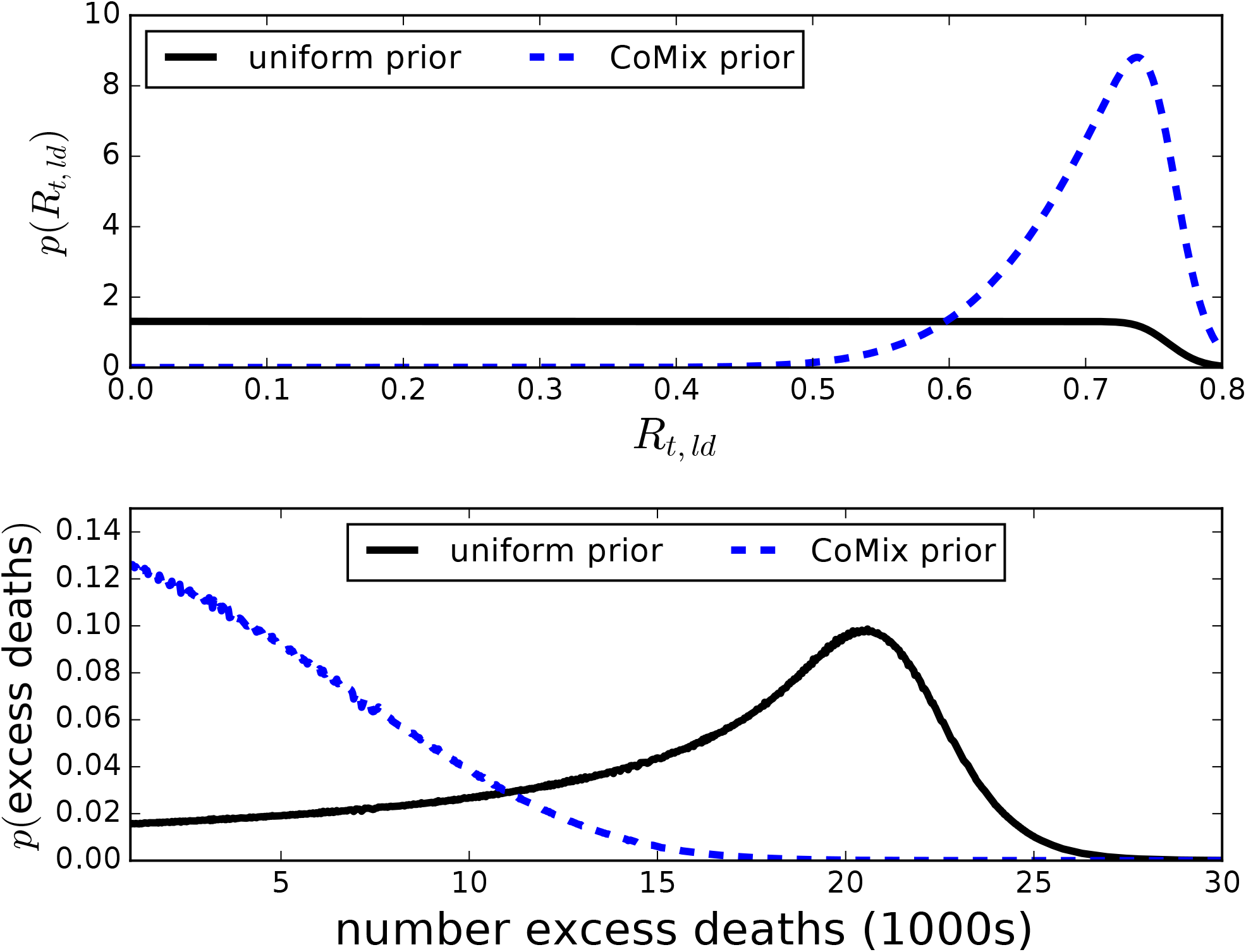
*Upper panel:* Posterior probability density for *R*_t,ld_, marginalised over *N*_*f*_, both for a uniform prior for *R*_t,ld_ (solid black line) and a prior based on the CoMix study (dashed blue line). Shown for *D*_*f*_ = 0.41 d. *Lower panel:* Probability density for excess number of deaths from fomites for *D*_*f*_ = 0.41 d over lockdown period, both for uniform and the CoMix prior on *R*_t,ld_.

The power-series solutions in Sec. 2.2 show fomites affect the death rates in second order. The magnitude of the departure from the prediction for a constant reproduction number may be seen from the effective reproduction number that would be inferred from the death rates assuming a counterfactual model with no fomites. Taking the exposure rate to be proportional to 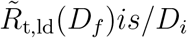, it follows from Eqs. (1) that 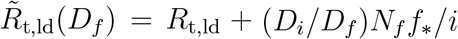. The effective reproduction numbers 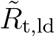 are computed from the solutions to models corresponding to the 95% confidence upper limits on *N*_*f*_ and their corresponding maximum likelihood values *R*_t,ld_ in Table 2 for the various values of fomite duration *D*_*f*_. Fig. 3 shows the fractional differences 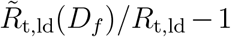, where *R*_t,ld_ is the maximum likelihood value for the reproduction number for a model with *N*_*f*_ = 0. The effective reproduction numbers become time-dependent, increasing with time by an amount that depends on *D*_*f*_. The fractional differences, however, are very small, exceeding 1% only when *D*_*f*_ is comparable to *D*_*i*_. The effect of fomites is too small to be discernable from the population statistics without additional information. Similarly, the mean excess numbers of weekly deaths for *N*_*f*_ *>* 0 compared with a counterfactual model having *N*_*f*_ = 0 are shown in the bottom panel of Fig. 1, along with the 68% confidence interval. The expected number of excess deaths over the lockdown period ranges from about 13000–16000 (with fewer excess deaths for large *D*_*f*_), corresponding to a fraction 0.6–0.7 of all deaths in this period. The probability distribution in the number of excess deaths, however, is very broad, as shown in the bottom panel of Fig. 2. No strong statement on the number of excess deaths resulting from fomite transmission may be made: any number up to all cannot be excluded at the 95% confidence level.

**Figure 3:**
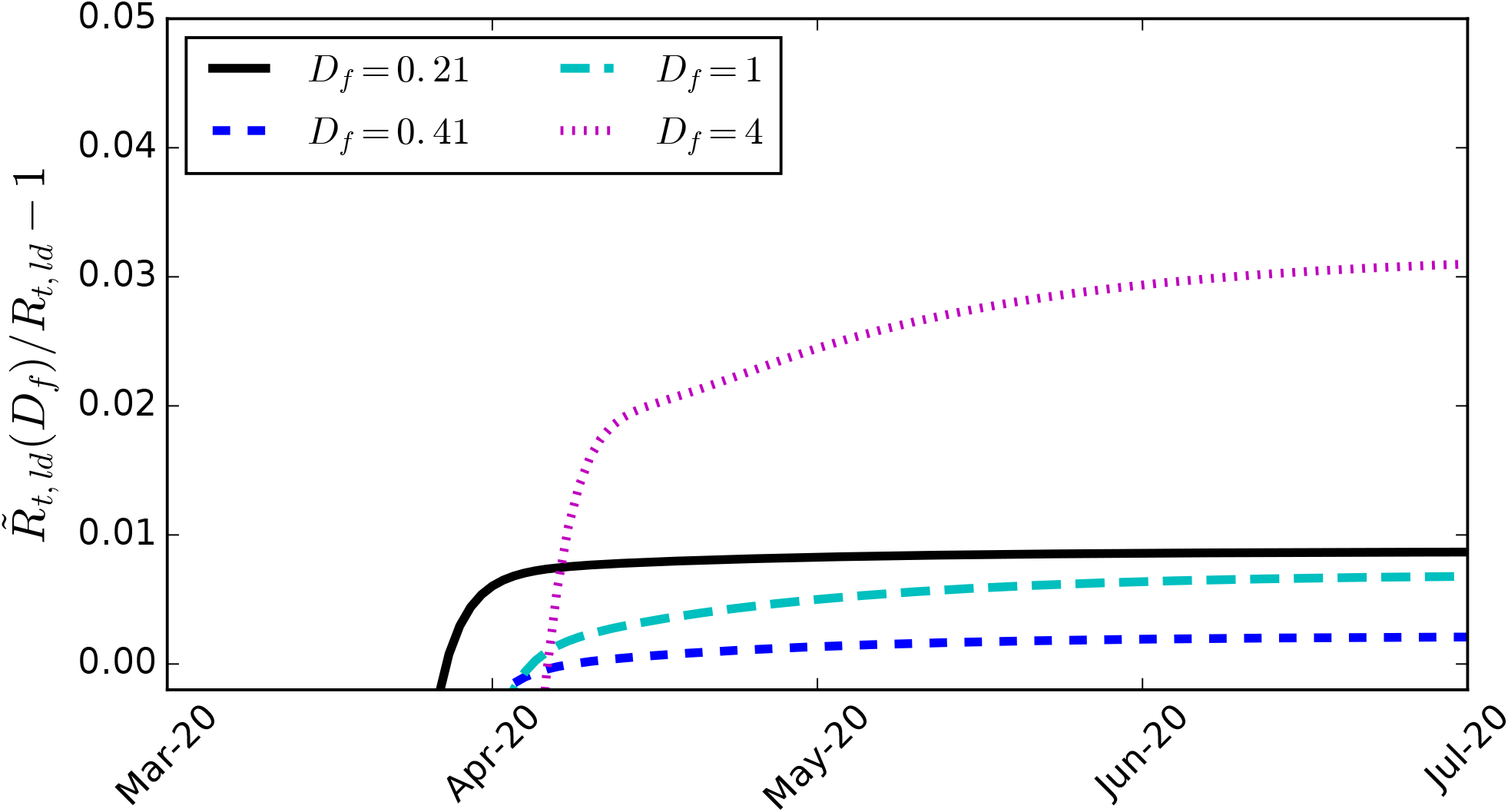
Relative difference between the effective reproduction number 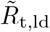 for models with *N*_*f*_ ≃ 0.14, *R*_t,ld_ ≃ 0.04, corresponding to the 95% confidence upper limits on *N*_*f*_, that would be inferred from the death rates assuming a counterfactual model with *N*_*f*_ = 0, compared with the maximum likelihood value for *R*_*t,ld*_ for a model with *N*_*f*_ = 0. Shown for various values of *D*_*f*_.

### 3.2. CoMix study prior

The CoMix study provides new information on the reduction in the number of social contacts per person during lockdown that limits the allowed range in the reproduction number in lockdown, breaking the degeneracy (co-linearity) between *R*_*t*_ and *N*_*f*_ in the effective reproduction number 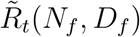. Allowing for fomites for which the transmission rate is constant, unchanged under lockdown conditions, the CoMix result on the reduction of social contacts before and after lockdown may be used to estimate a prior probability distribution on the reproduction number in lockdown, *R*_r,ld_, as follows. The basic reproduction number found before lockdown is re-interpreted as an effective basic reproduction number 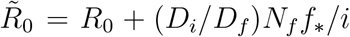. For any given values of *N*_*f*_ and *D*_*f*_, the value of 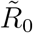 at the moment of lockdown, at time *t*_ld_, is used to provide an estimate for the actual basic reproduction number in the model, 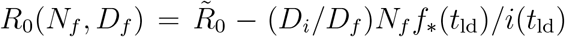. Under the assumption that the reproduction numbers are directly proportional to the number of social contacts, for any given reproduction number *R*_t,ld_ during lockdown, the likelihood for the model is multiplied by the likelihood of the ratio *f*_ld_ = *R*_t,ld_*/R*_0_(*N*_*f*_, *D*_*f*_), taken to be proportional to a normal distribution with mean 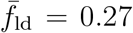 and standard deviation 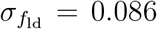, as inferred from Jarvis et al. (2020). (It is noted in practice it is sufficient to use the small *D*_*f*_ limit for obtaining 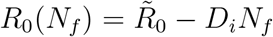, as the difference in estimators is small compared with *R*_0_.)

The probability distribution for *R*_t,ld_, marginalised over *N*_*f*_, is shown in Fig. 2 for *D*_*f*_ = 0.41 d. The results are nearly independent of *D*_*f*_ over 0.21 ≤ *D*_*f*_≤4 d. In contrast to models with a uniform prior on *R*_t,ld_, the CoMix study result on the reduction of social contacts in lockdown corresponds to a maximum likelihood value for the reproduction number during lockdown of 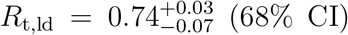, after marginalising over *N*_*f*_. This may be compared with the estimate for a model with *N*_*f*_ = 0 of *R*_t,ld_ = 0.76±0.02 (68% CI), so that allowing for fomites little affects the expected value but broadens the confidence interval.

By contrast, the marginalised probability distribution for *N*_*f*_ continues to be flat, peaking at *N*_*f*_ = 0. The marginalised 95% upper limit on *N*_*f*_ is *N*_*f*_ ≲ 0.035 d^−1^ infectious person^−1^ for 0.21≤*D*_*f*_≤1 d and *N*_*f*_ *<* 0.028 d^−1^ infectious person^−1^ for *D*_*f*_ = 4 d. Corresponding to these limits on *N*_*f*_, the maximum likelihood value for the reproduction number during lock-down is *R*_t,ld_ = 0.6. Marginalised over *N*_*f*_, *R*_t,ld_ *<* 0.77 at 95% confidence. The corresponding maximum likelihood values for *N*_*f*_ vanish for all values of *D*_*f*_. The overall maximum likelihood solution corresponds to *N*_*f*_ = 0 and *R*_t,ld_ = 0.76 for all values of 0.21≤*D*_*f*_, ≤4 d. Consequently, conservatively no fomite signal is detected, but a much more restrictive upper limit on the fomite rate may be set compared with the case of a uniform prior on *R*_t,ld_.

The mean weekly death rate for *D*_*f*_ = 0.41 d is shown in the upper panel of Fig. 4, along with the 68% confidence interval. The lower panel shows the mean excess numbers of weekly deaths for *N*_*f*_ *>* 0 compared with a counterfactual model having *N*_*f*_ = 0, along with the 95% upper limit. Summing over the shown lockdown period gives an expectation of about 4960 excess deaths, compared with about 22900 total deaths. This corresponds to an expected fractional death excess of 0.22. The probability density for the total number of excess deaths over the lockdown period is broad, but peaks at a lower number of excess deaths compared with a uniform prior on *R*_t,ld_, as shown in the lower panel of Fig. 2. The 95% upper limit on the number of excess deaths is 12000, corresponding to a fraction 0.52 of all deaths. This is typified by a model for *D*_*f*_ = 0.41 d with *N*_*f*_ = 0.035 d^−1^ infectious person^−1^ and *R*_t,ld_ = 0.59 (see Table 3), for which the fraction of excess deaths from fomites during lockdown is 0.51. Similar results are found for *D*_*f*_ = 0.21 and 1 d. The number of excess deaths for *D*_*f*_ = 4 d, however, is somewhat smaller. The expected number of excess deaths is about 4200 over the lockdown period, corresponding to a fraction 0.18 of all deaths during this time, with an upper limit of 10300 excess deaths (95% CL), corresponding to a fraction 0.45 of all deaths during lockdown.

**Table 3:** Model results for a uniform prior for *N*_*f*_ and a prior for *R*_t,ld_ based on the CoMix study. The columns are: (1) fomite duration (days); (2) 95% upper limit on *N*_*f*_ (per infectious person per day) and (3) the corresponding maximum likelihood value for *R*_t,ld_; (4) 95% upper limit on *R*_t,ld_ and (5) the corresponding maximum likelihood value for *N*_*f*_ (per infectious person per day).

| $D_f$ | $N_f$ (95% ul) | $R_{t,ld}$ (ml) | $R_{t,ld}$ (95% ul) | $N_f$ (ml) |
| --- | --- | --- | --- | --- |
| d | d<sup>-1</sup> |  |  | d<sup>-1</sup> |
| 0.21 | 0.034 | 0.59 | 0.77 | 0 |
| 0.41 | 0.035 | 0.59 | 0.77 | 0 |
| 1 | 0.034 | 0.58 | 0.77 | 0 |
| 4 | 0.028 | 0.61 | 0.77 | 0 |

**Figure 4:**
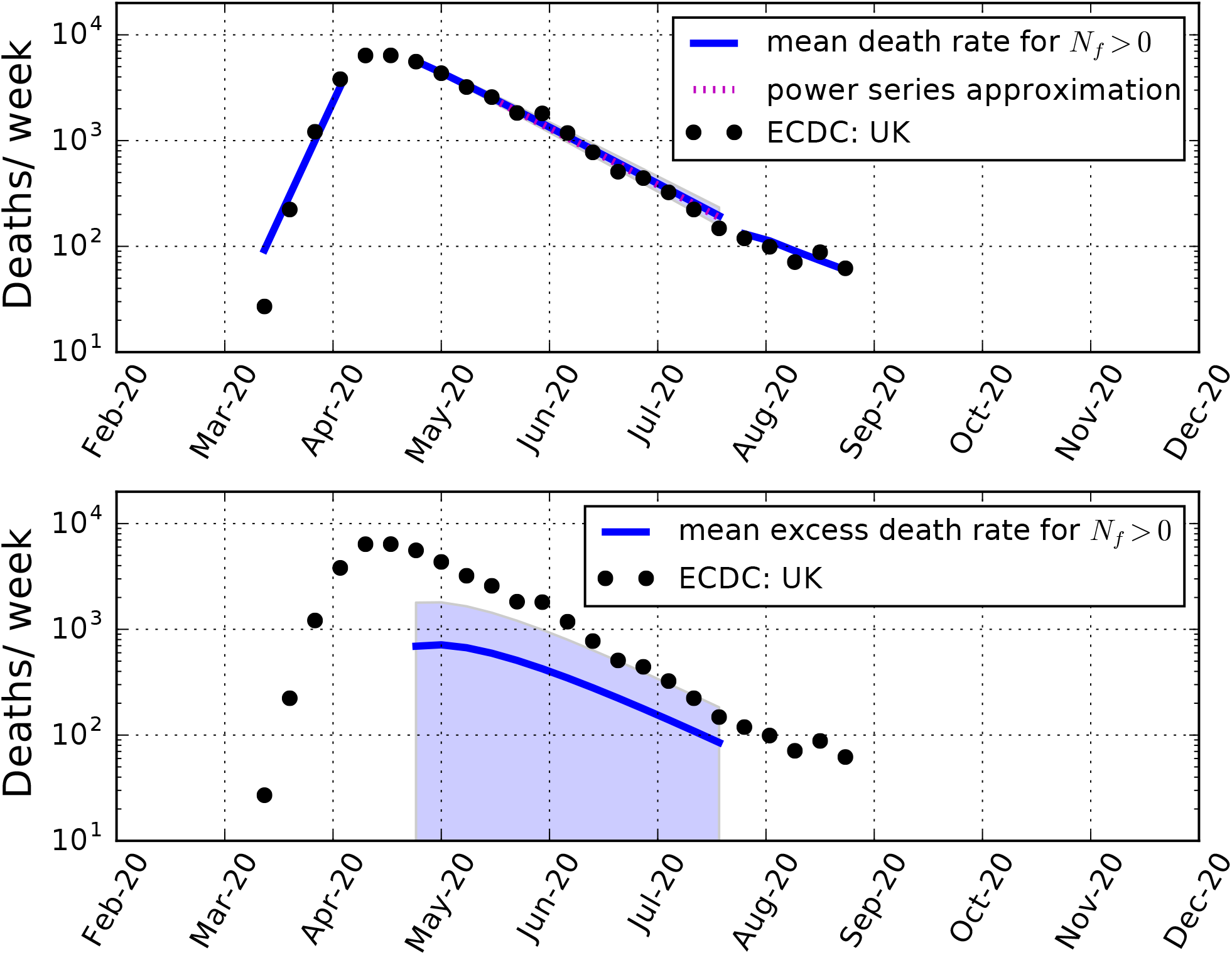
*Upper panel:* Weekly death rates in the UK. Black data points: the rates reported by the ECDC. Solid blue lines: Mean weekly deaths predicted for *D*_*f*_ = 0.41 d and assuming a uniform prior for *N*_*f*_ and a prior for *R*_t,ld_ after lockdown based on the CoMix study. Also shown is the 68% range of uncertainty in the mean predicted number of deaths during lockdown (shaded region). Red dotted line: Power series approximate solution during the lockdown period (see Appendix). *Bottom panel:* Blue solid line: Mean predicted excess deaths per week arising from fomites, along with the 95% upper limit (shaded region).

## 4. Discussion

### 4.1. Parameter constraints

Fomites are recognised as potential contributors to the transmission of the novel coronavirus SARS-CoV-2, however statistically quantifying the contribution is rendered difficult by the rarity of fomites in the general environment. During a long lockdown period, the reproduction number from direct transmission may be nearly constant, simplifying the evolution of the infection. Population statistics are examined as a possible means to constrain the fomite transmission rate *N*_*f*_ during a lockdown. Fomite transmission alters the evolution in the rate of deaths compared with a non-fomite model with a constant reproduction number, although the difference is small. Adding a fomite contribution is not found to improve the likelihood during the lockdown phase allowing for uniform priors on the fomite transmission rate *N*_*f*_ and reproduction number for direct transmission during lockdown *R*_t,ld_. Upper limits are placed of *N*_*f*_ *<* (0.13−0.14) d^−1^ infectious person^−1^ (decreasing with *D*_*f*_) and *R*_t,ld_ *<* 0.72−73 (increasing with *D*_*f*_) (95% CL). The joint likelihood function for both *N*_*f*_ and *R*_t,ld_ has a sharp ridge at *R*_t,ld_+*D*_*i*_*N*_*f*_≈0.76, expressing their near statistical co-linearity. Whilst the formal maximum likelihood model corresponds to *N*_*f*_ = 0, the probability distribution for the number of excess deaths is broad. The expected number of excess deaths arising from fomites is 0.6–0.7 of the total during lockdown (decreasing with *D*_*f*_), and the possibility that all deaths arose from fomites may not be excluded at better than 95–98% confidence (increasing with *D*_*f*_).

Including the CoMix study results on the reduction in social contacts between people in the UK following the near complete lockdown imposed in March 2020 modifies the prior distribution on the reproduction number, allowing a tighter constraint to be placed on the fomite transmission rate of *N*_*f*_ ≲ 0.03 d^−1^ infectious person^−1^ (95% CL) for 0.21 *< D*_*f*_ *<* 4 d. The reproduction number during lockdown is found to be 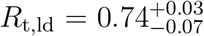 (68% CI), after marginalising over *N*_*f*_. The probability distribution for *R*_t,ld_ is little sensitive to the addition of fomites, so that models without fomites should provide reliable estimates even if fomites are active. The value found here is consistent with other estimates, but on the lower end of the range (DHSC/SAGE, 2021).

To give the limits on *N*_*f*_ some context, the mean number *C*_*f*_ of potentially contaminatable objects a person may come into contact with per day must be specified. An estimate for the post is *C*_*f*_≃0.57 d^−1^ person^−1^ (Meiksin, 2020). For food items, the Office for National Statistics estimates an average spend of £63.80 per week for an average household (ONS, 2020) of 2.4 members (ONS, 2021). For an average product value of £2, this corresponds to *C*_*f*_ ∼ 2.5 d^−1^ person^−1^ for post and food items combined. The upper limit allowing for the CoMix prior then corresponds to a fomite transmittivity upper limit of *T*_*f*_ *<* 0.014 (95% CL), or at most about 1 in 70 objects that comes into contact with an infectious person transmits the infection to a susceptible person.

Quantifying the impact of fomites in terms of the excess deaths compared with a counterfactual non-fomite model, the expected number of excess deaths during lockdown is found to be 22% of all deaths during this period. The probability distribution on the number of excess deaths is found to be broad, however, so that the 95% upper limit on the number of excess deaths is 52% of all deaths during lockdown for 0.2≤*D*_*f*_≤1 d. For fomites with a duration time comparable to the infectious period, the limits are somewhat more restrictive. For *D*_*f*_ = 4 d, the expected number of excess deaths is 0.18 of all deaths during lockdown, with a 95% upper limit of 0.45. Regardless of fomite duration time, allowing for the CoMix study prior on the reproduction number during lockdown shows that it is highly unlikely that most deaths from COVID-19 were caused by transmission through lockdown-independent fomites.

### 4.2. Limitations of study

Placing constraints on the contribution of fomites during lockdown requires some key assumptions. In this context, fomites may be divided into two types, those that scale like the number of social contacts and those independent of the number of social contacts. Only the latter, such as fomites arising from essential services that continued through the lockdown like post deliveries and food and medicine purchases, may be constrained using the change in the evolution of the population statistics following lockdown. Fomite transmission rates that scale with the number of social contacts, such as may occur for fomites in office spaces, likely scale with the reproduction number for direct transmission, and so their effects on population evolution statistics are indistinguishable from those of direct transmission. It is not possible to rule out from population statistics the possibility that most of the transmission prior to lockdown arose from workplace fomites. Although contamination through household fomites may have then been expected to increase during lockdown, an increase in fomite transmission in a domestic setting may have been masked by the limited number of additional family members that could have been infected. Increased hand-washing in the home could in principle also be a factor.

The strongest constraints on fomite transmission in this study rely on the measured change in the number of social contacts following lockdown from the CoMix study in the UK. It is assumed the change in the reproduction number is proportional to the change in the number of social contacts, and that the proportionality factor is independent of age. The measured reduction may be subject to several biases (Jarvis et al., 2020): there is a possibility of recall bias, as the study requested information about the previous day; the sample may be subject to selection bias if preferably people observing the lockdown replied to the survey; also children were not interviewed, so that child-child contacts were inferred from the POLYMOD survey.

The maximum likelihood models with fomites were found to result in a small increase in the transmission with time compared with the maximum likelihood models without fomites. This difference accounts for the more stringent upper limits on the fomite contribution for longer fomite duration times. The increase may be interpreted as evidence for a slightly increasing reproduction number with time, in which case the fomite contribution would be even smaller. The alternative of a decreasing reproduction number with a larger fomite contribution than the upper limits found here cannot be ruled out, but it would seem unlikely the reproduction number would decrease in anticipation of an easing of the lockdown, and in any case this possibility would have only a small effect on the limits found.

### 4.3. Suggestions for further progress

CoMix is part of a European partnership involving 17 countries^2^. As more results on the change in social contacts are published, similar analyses to this may be performed to tighten constraints on fomite transmission of the SARS-CoV-2 virus within Europe.

The high availability of tests for infections may now allow more direct estimates of transmission through post and grocery deliveries by correlating illnesses between recipients and deliverers, including sorters and stockers, who may have been in an asymptomatic infectious phase when handling items. Such correlations may be made possible with the assistance of data from the track-and-trace smartphone app system ^3^ in conjunction with appropriate data protection measures.

## 5. Conclusions

Because of their longevity on surfaces, fomites alter the time evolution of populations in an epidemic compared with direct transmission alone. The differences, however, are too small to detect when the fomite duration is shorter than the duration of the infectious phase. For uniform priors on the reproduction rate and fomite transmission rate, only coarse upper limits may be set on the fomite transmission rate. Allowing for a measurement of the change in the number of social contacts when a lockdown is implemented, as provided by the CoMix study, and under the assumption that the reproduction number scales with the number of social contacts, more restrictive constraints may be placed on the role of fomites. Using data for the UK from the lockdown in March to July 2020, it is found that fomites that act independently of a lockdown, such as delivered post or grocery items, contributed to fewer than about half of the total deaths from COVID-19 during the lockdown (95% CL), and likely fewer than a quarter.

## Data Availability

No new data were generated in this research.

## Declaration of Competing Interests

None.

## Funding

This research did not receive any specific grant from funding agencies in the public, commercial, or not-for-profit sectors.

## Appendix A.

Power-series solutions of Eqs. (1) in *r* are constructed by first dividing the equations by *dr/dt* = *i/D*_*i*_ to transform them to

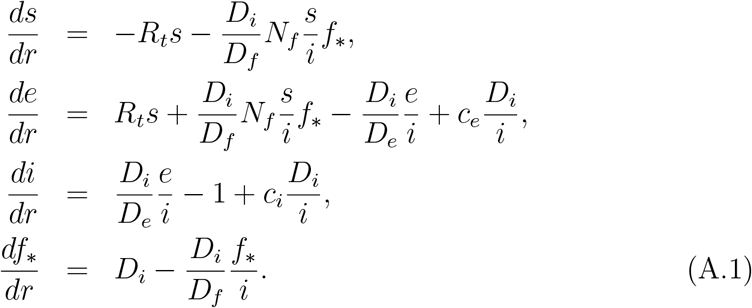

Power-series solutions around *r* = *r*_0_ = *r*(*t*_0_) after a time *t*_0_ are assumed:

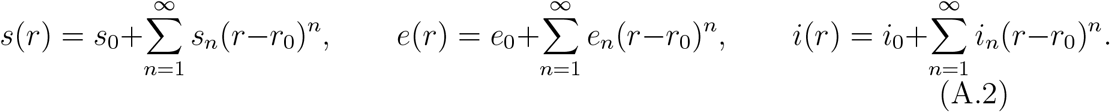

It is numerically convenient to use the integrated solution for the fomite term:

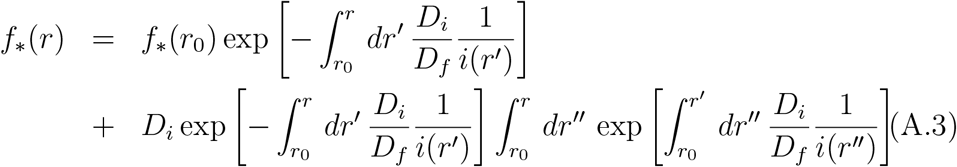

To second order, the coefficients are

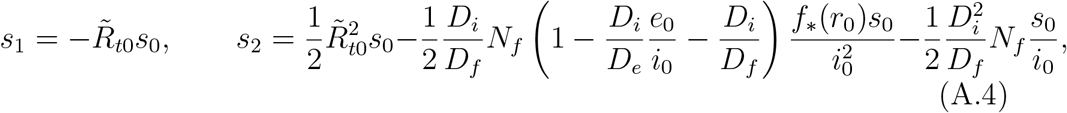

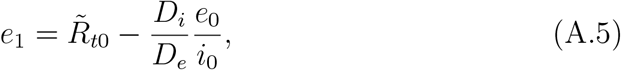

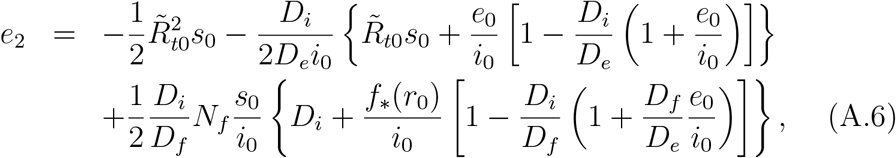

and

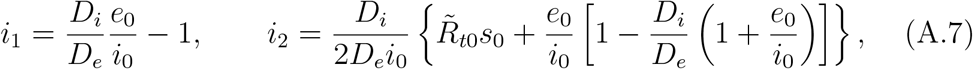

where the effective reproduction number 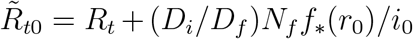 has been defined.

The forms of the coefficients show that to first order, the fomite term does not affect the evolution of the populations other than through a re-scaling of *R*_*t*_ to 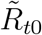. At second order, the evolution of the susceptible and exposed populations show additional contributions from the fomites that are not eliminated by a re-scaling, whilst the evolution of the infectious population, in contrast, may still be re-scaled to second order. Using the expression for *i* to second order, Eq. (A.3) becomes

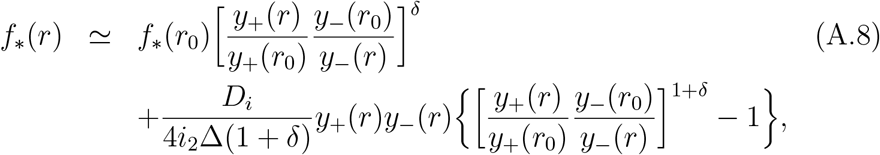

where *y*_*±*_(*r*) = Δ ± [*i*_1_ + 2*i*_2_(*r* − *r*_0_)], 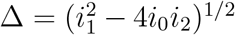 and *δ* = *D*_*i*_*/*(*D*_*f*_ Δ). (For applications here, 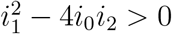.)

The evolution of the populations may be expressed as a function of *t* through

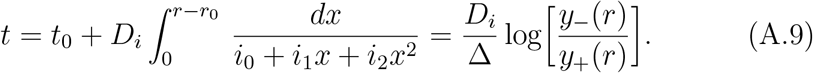

An example solution is illustrated in the upper panel of Fig. 4 for a solution at the 98% upper limit of *N*_*f*_ = 0.042 allowing for the CoMix prior, and the corresponding maximum likelihood value *R*_t,ld_ = 0.55. The predicted number of deaths agrees with the numerical solution for these parameter values to within 1%.

## Footnotes

1 https://www.ecdc.europa.eu/en/covid-19/data-collection

2 https://www.uhasselt.be/UH/71795-start/The-CoMix-study

3 https://www.nhs.uk/apps-library/nhs-covid-19/; https://protect.scot

## Notes

### Competing Interest Statement

The authors have declared no competing interest.

